# A Pilot Feasibility Study of PAL-CHW-PDAC, a Digitally Enhanced CHW-led Intervention to Facilitate Stepped Palliative Care in Patients with Pancreatic Cancer

**DOI:** 10.1101/2025.10.06.25337465

**Authors:** Nikhil R. Thiruvengadam, Maud Celestin, Lizbeth Rivas, Nishita Matangi, Arman Bahmani, Matthew Oroso, Joel Brothers, Kendrick Che, Andrew Chang, Emmanuel Eguia, Paul Leonor, Gina Mohr, Raja Narayan, Sagar Pathak, Susanne Montgomery, Betty Ferrell

## Abstract

**Background:** Pancreatic Ductal Adenocarcinoma (PDAC) is associated with substantial morbidity and poor quality of life (QOL). Early palliative care (EPC) has been shown to improve quality of life and reduce symptom burden in PDAC, but only a minority of PDAC patients receive it. Using three conceptual models, we developed a CHW-led intervention to help facilitate stepped PC.

**Objectives:** To evaluate the feasibility, acceptability, and preliminary effectiveness of PAL-CHW-PDAC.

**Design:** A single-center prospective single-arm pilot study

**Setting/Subjects:** Newly-diagnosed PDAC patients (n=48) were included along with their family caregivers (FCGs)

**Measurements:** Acceptability and Feasibility were defined using the RE-AIM framework and using the Acceptability of Intervention Measure. Secondary outcomes were symptom burden, quality of life, attitudes towards PC, and ACP completion.

**Results:** The median age was 69. Of those, 29 (60%) were Hispanic, 25 (52%) had metastatic PDAC, while 23 (48%) had borderline resectable/resectable PDAC at diagnosis. Four patients died before completion of the intervention, while all remaining patients completed it. Regarding acceptability, 94% of patients strongly agreed that the intervention was acceptable. Themed common responses highlighted by patients centered around the critically important role of the CHW in the process, including in appointment and referral scheduling, especially as symptoms worsened. Patient’s physical HRQoL and symptom burden improved from baseline to 12 weeks, while 83% of patients completed ACP.

**Conclusions:** The PAL-CHW-PDAC intervention was feasible and acceptable to PDAC patients and improved symptom burden. Prospective clinical trials are needed to assess the intervention’s ability to address the QOL needs of PDAC patients.

## Background

Pancreatic Ductal Adenocarcinoma (PDAC) is projected to become the 2nd largest contributor to cancer mortality by 2030.^1^ PDAC also carries significant morbidity, with 72% of PDAC patients having a significant decrease in health-related QoL (HRQoL) in the 30 days following diagnosis.^2, 3^ Early Palliative Care (PC) within 2-4 weeks of diagnosis has been previously shown to improve HRQoL.^4, 5^ However, only as few as 30% of patients with PDAC receive specialty PC services, as referral and availability delays often make them unavailable for this fast-progressing illness.^6–10^ Indeed, most patients with PDAC receive PC care only in the last month of life, limiting the impact of PC on mitigating early PDAC-related morbidity.^10–12^ Reasons for low PC utilization include a shortage of specialty PC providers, a clinical paradigm where PC referrals are placed only after establishing care with oncology, a lack of awareness of, and misconceptions about its benefits among patients.^6, 13, 14^

Multiple models have been studied to improve PC uptake among patients with advanced cancer. Stepped PC models have been developed in which visits are “stepped up” as the disease or symptoms worsen. A recent RCT demonstrated that stepped PC was non-inferior to early scheduled PC in advanced lung cancer, with regard to QOL, required fewer PC visits and was more scalable.^15^ Furthermore, a multi-center RCT demonstrated that telehealth-based EPC was non-inferior to in-person care for delivering EPC in lung cancer.^16^ However, these interventions have not been studied in PDAC. The older age of patients with PDAC (median age 73), low digital literacy levels and rapid deterioration have made using telehealth challenging.^17^ Community health workers (CHWs) are frontline, public health workers and trusted members of the community served,^18^ who help bridge the gap between individuals and healthcare systems. Interventions facilitated by CHWs have been found to reduce acute care costs and improve ACP but there is limited data for PDAC.^19^

To address this gap, we collected qualitative feedback from patients and family caregivers (FCGs)about their experience and then used it to develop a digitally enhanced CHW-led intervention, PAL-CHW-PDAC focused on symptom burden.^20^ The intervention facilitates stepped PC, including symptom management, care navigation, and patient education, paired with an SMS-based digital application that assesses patients’ symptom burden weekly via patient-reported outcomes (PROs). Our objective in this study was to assess the acceptability and feasibility of PAL-CHW-PDAC in a prospective single-arm study (NIH stage 1).

## Methods

### Study Setting and Participants

We conducted a prospective single-arm feasibility study of PAL-CHW-PDAC at Loma Linda University Cancer Center. The local institutional review board approved the study (IRB# 5240144), as did the Office for Human Research Oversight (Protocol Number E05835.2a, the study’s funder). The study authors had access to the data and reviewed and approved the final manuscript. The inclusion criteria were as follows: age > 18 and newly diagnosed PDAC (within 2 weeks of histologic diagnosis). Caregivers were allowed to participate in the intervention, but for the purpose of this study, patients were considered the research subjects.

### Intervention Development

We used a multiphase study with formative (Phase 1) and summative (Phase 2) evaluations, guided by the NIH Stage Model^21^ for the development of complex behavioral interventions. (We advanced from NIH Stage 0 (Basic Research involving literature review and key-informant interviews (KIIs) with patients, FCGs, and providers) to Stage 1 (*Intervention Generation/Refinement*/Pilot Testing). We followed a human-centered design approach, engaging a diverse set of key stakeholders at every step of this intervention’s development. Figure 1 describes the details of our intervention development

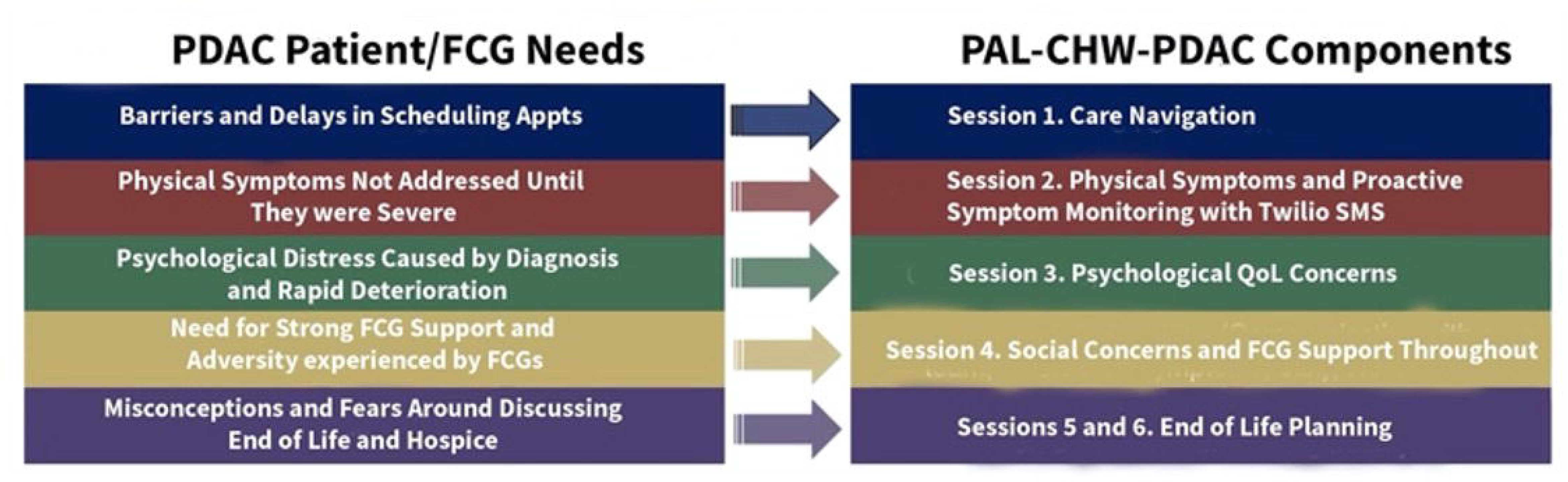

### Conceptual Frameworks

Our intervention is informed by 3 conceptual models: 1) The QOL model (physical, psychological, social, and spiritual well-being)^22, 23^ developed by Ferrell et al. describing the needs of patients and FCGs, 2) the 8 domains from the National Consensus Project for Quality PC, informing the content and structure of the intervention^24^, and 3) the Social Ecological Model (SEM)^25^ to conceptualize the multi-level causes of health outcomes. Therefore, PAL-CHW-PDAC addresses community capacity at the individual, interpersonal, organizational, and community levels.

### Phase 1 Formative Evaluation (NIH Stage 0)

The intervention was developed based on a literature review, existing PDAC educational materials, and key informant interviews (10 patients diagnosed with PDACs and 11 caregivers) from the local community.^26^ Themes revealed specific challenges related to the medical journey and treatment, barriers to navigating the healthcare system, and resistance to end-of-life conversations, planning, and care. Suggestions for healthcare providers were also gathered from these interviews. These themes guided the final design of the intervention in content and delivery modality.

### Phase 1 Interventional Development (NIH Stage 1a)

In Figure 2, we demonstrate the needs of patients/FCGs that were mapped to the intervention. The intervention creation was described previously. We created a standardized training program for the CHWs. As part of the intervention, we developed a 3-week training manual and curriculum for the CHW to orient them to this patient population and the intervention, followed by 1 week of practice teaching, simulation, and observation. This training involves lectures from gastroenterologists, surgeons, oncologists, PC physicians, nurses, social workers, and nutritionists; attending tumor board, new patient conference, and PC rounds to gain exposure to the care patients with PDAC receive. At the end of this training, the CHWs had 1 week to conduct practice visits, supervised by Dr. Celestin, to ensure CHWs deliver the intervention with fidelity to the training manual

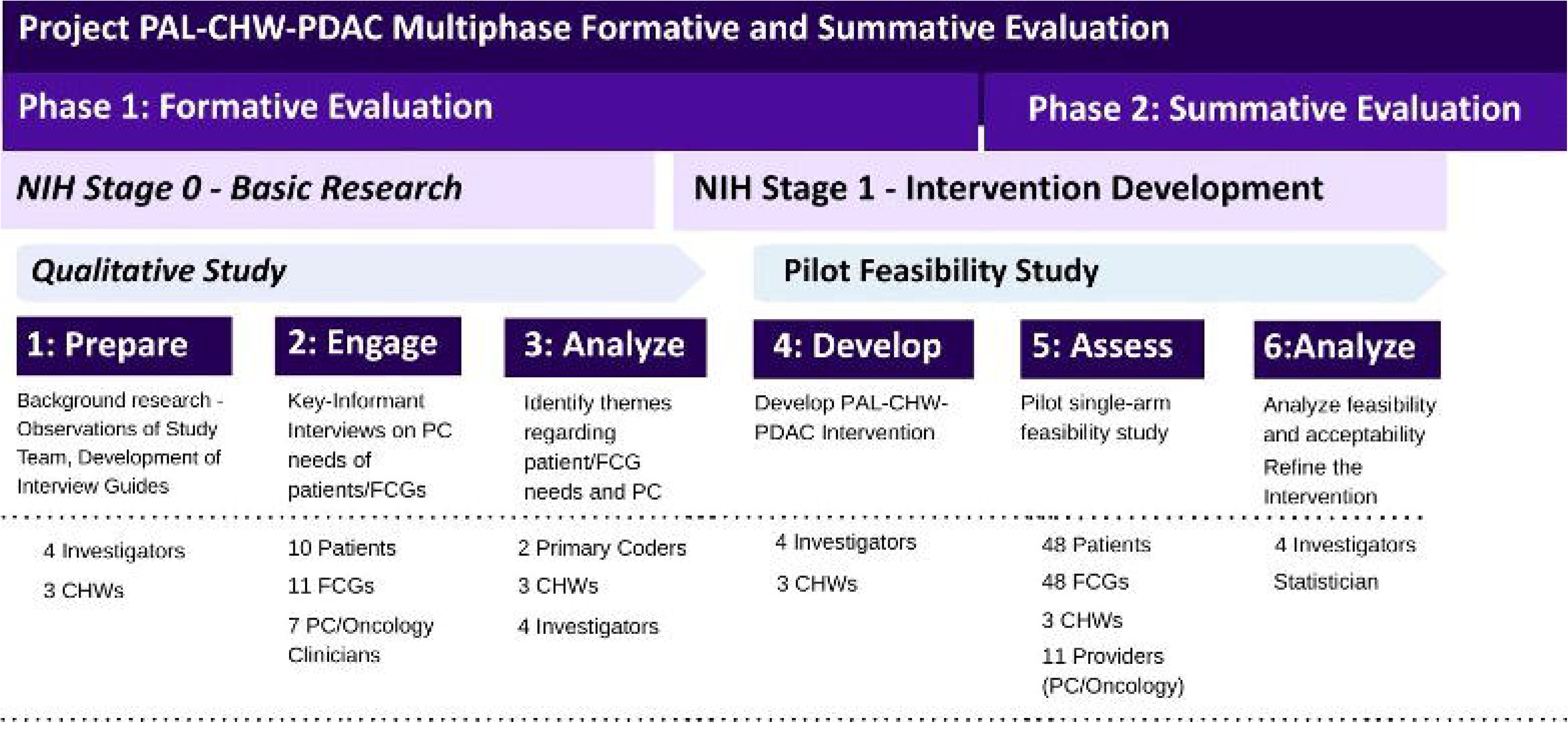

### Intervention timeline

The intervention is to be delivered in four weekly visits during the first four weeks after enrollment, by a CHW either through in-person visits (in hospital or patient homes) or via Zoom or telephone. There are two additional monthly visits in months 2 and 3. The intervention is designed to engage both the patient and caregiver but can be delivered to a patient alone. The CHW also conducted optional follow-up sessions as needed, including a dedicated session on explaining the role of hospice based on the patient’s needs. Participants received a $10 gift card after completing each session, and one for answering each of the three surveys (at baseline, at 4 weeks, and at 12 weeks).

Patients with newly diagnosed PDAC were approached by the study investigators; the study and its risks were explained, and if the patient was willing to participate, informed consent was obtained. Once enrolled, they were asked to complete the baseline instruments, and in the first week after enrollment, were introduced to the CHW, either in the clinic or by video call. During the patient’s introduction to the CHW, it was also determined whether the first visit would be at home or via Zoom, whether the patient’s FCG was available, and whether both the patient and the FCG would be invited to attend the intervention. Table 1 details the contents of each session with the CHW.

**Table 1.**
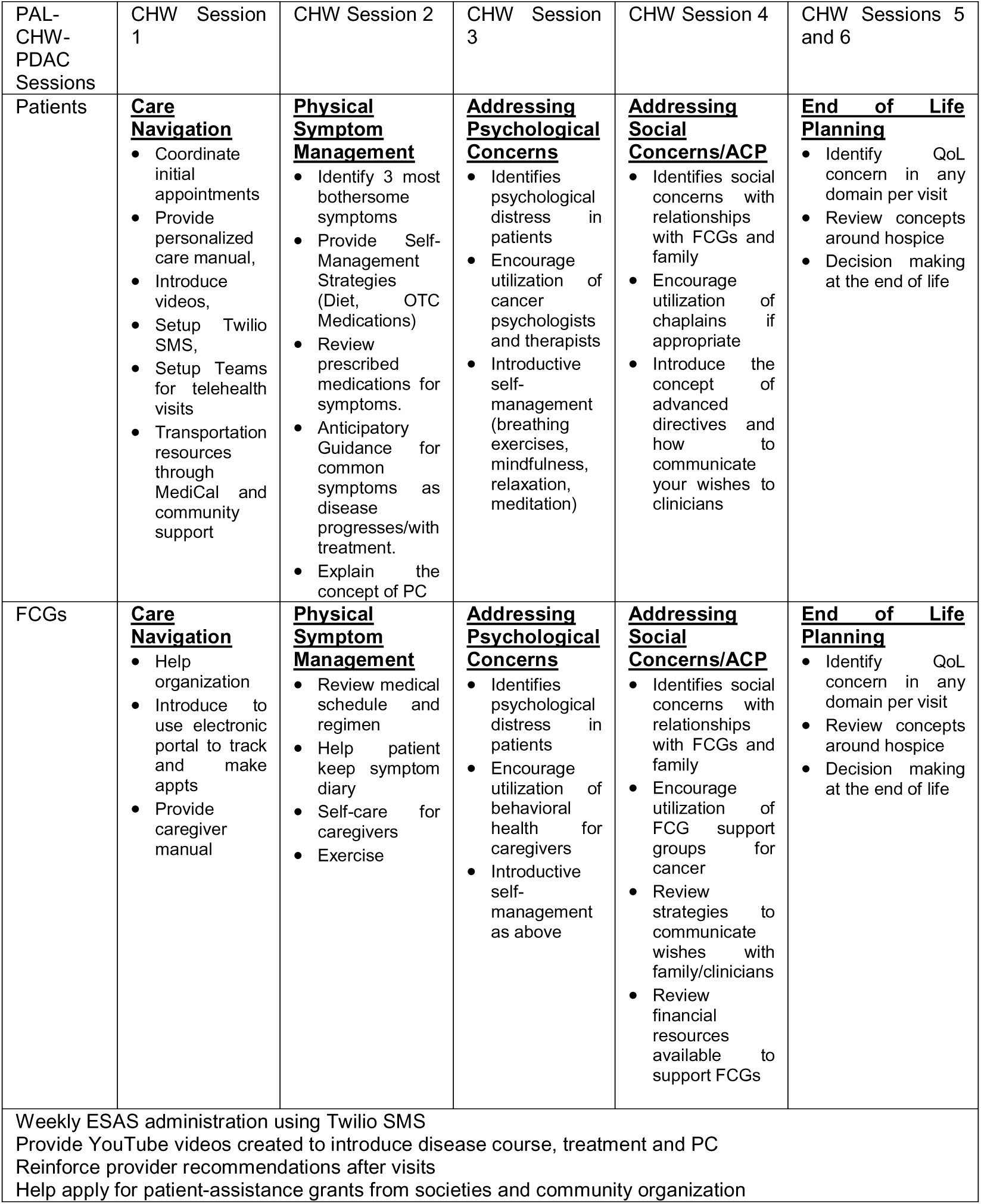
Contents of the PAL-CHW-PDAC intervention.

CHWs also provided patients and caregivers with written educational resources covered during the personalized CHW-led session discussion, including a wire-bound manual on PDAC and its treatment, emphasizing symptom recognition and quality of life. Since it is well documented that information retention is low at the time of diagnosis, manual content was developed in English and Spanish with clearly marked inserts that explained terms the caregiving team might use, but patients may not fully understand. In recognition of the severity of the content and different learning styles, we delivered the intervention in three complementary modalities: in person, in writing, and as CHW narrated videos—all covering the same content, thus allowing patients and their caregivers to revisit the content as often as they needed in the patient’s choice of language (English or Spanish).

### Digital Component of the Intervention

During that first visit, the CHW also ensured that the patient could access and use the REDCap Twilio short message service, which sent the patient the Edmonton Symptom Assessment Scale^27^ on a weekly basis, along with appointment reminders. They also helped set up the patient to use the MyChart or Teams platforms for telehealth visits.

### Proactive Symptom Monitoring and the Stepped PC Component

Each week, patients received the ESAS via SMS through Twilio to assess the severity of 7 symptoms, and the CHW reviewed their responses within 24 hours of completion. (Table 2) If a patient reported any symptom with intensity >3, the CHW called the patient to reinforce the clinical team’s existing recommendations for managing their symptoms and self-management strategies. (Table 3) The CHW also coached the patient on how to communicate their symptom concerns to their clinical team, either through the MyChart portal or by calling their provider. While all patients received a PC consultation at diagnosis and an initial visit, additional visits in the intervention arm were triggered if a patient reported symptom intensity greater than 7/10 on the weekly ESAS, experiences disease progression, or had other QOL concerns. If the patient reported any serious concerns in any of the 4 QOL domains, the CHW will escalate contact with the medical team as appropriate

**Table 2.** Weekly Proactive Symptom Monitoring with the ESAS administered over Twilio SMS.

| If the ESAS is the following: |  |
| --- | --- |
| Symptom Score 1-3 | CHW reinforces self-management strategies and reinforces prior provider recommendations at next visit. |
| Symptom Score 4-6 | First, CHW will call the pt to elicit more information and see if the symptom can be resolved with the current plan. If it is new, the CHW will encourage the pt to contact their oncologist's office to report it. The CHW will check back in 24 hrs. to ensure that the pt was contacted by their Oncology team and the symptoms are improving. |
| Symptom Score > 7 | The CHW will contact the Oncology RN who will contact patient to discuss further and help setup an expedited visit with the oncologist or PC team to discuss this symptom. If symptom has been previously addressed the CHW will reinforce provider recommendations for its management. |

**Table 3.** Outcome Measures for Pilot Feasibility Study.

| Primary Outcome | Measure | Definition |  |  |
| --- | --- | --- | --- | --- |
| Feasibility and Acceptability | Reach | Quantitative<br>1. Proportion of patients who participate in the study<br>2. Percent who complete exit interview |  |  |
|  | Effectiveness | Quantitative<br>1. Percent attrition after enrollment<br>2. Acceptability of – Measure/Intervention Appropriateness Measure/Feasibility of Intervention Measure (AIM-IAM-FIM score) |  |  |
|  | Adoption | Qualitative (interviews with Patients, FCGs, Providers)<br>1. Were there any unintended effects of the intervention (positive or negative)?<br>2. Was the intervention helpful to patients, FCGs, and providers. |  |  |
|  | Implementation | Quantitative<br>1. Proportion of patients who participate in the study<br>Qualitative (Interviews with Patients and CHWs)<br>1. Can this intervention be implemented in clinical practice |  |  |
|  | Maintenance | Qualitative (Interviews with Providers and CHWs)<br>1. Can intervention delivery be maintained by the CHW?<br>2. Did the intervention add to physician workload or burden? |  |  |
| Secondary Outcome | Measures | Baseline | 4 wk | 12 wks |
| Symptom Burden | The National Comprehensive Cancer Network Functional Assessment of Cancer Therapy (NCCN-FACT) Hepatobiliary-Pancreatic Symptom Index (NFHSI)<sup>40</sup> (Ranges from 0 to 72 with higher being better) | X | X | X |
| QOL | FACT-Hep<sup>40</sup> (Ranges from 0 to 180 with higher being better) | X | X | X |
| Psychological Distress | NCCN Distress Thermometer<sup>41</sup> | X | X | X |
| Attitudes Towards Palliative Care | Palliative Care Attitudes Survey (PCAS) 9 | X | X | X |
| Attitudes towards ACP | ACP Engagement Survey<sup>42</sup> | X | X | X |
| Resource Utilization | Telehealth Utilization, Hospice Utilization, Nutritionist utilization, ER visits, hospitalizations |  |  | X |

#### Study Outcomes

Upon enrollment, participants were administered baseline surveys to determine demographic characteristics and assess symptoms and attitudes towards PC. The primary and secondary outcomes for this study are seen in Table 3.

#### Quantitative Analysis

Categorical data are presented as frequencies and percentages (Table 1). Continuous data are presented as the mean with standard deviation or median with interquartile range (IQR) according to distribution. To assess a preliminary pre-/post-effect, we compared survey data between baseline and 4 weeks using generalized linear models to compute contrasts for continuous variables from the FACT-Hep subscores, PCAS-9, and ACP Engagement Survey scores. We used an independent *t-*test to compare acceptability and feasibility rates between English and Spanish-speaking patients.

#### Qualitative Analysis

At the end of the study, key informant interviews (KIIs) were conducted with each patient and CHW, and thematic analyses were conducted to compare themes by gender and stage. The KIIs were audio recorded, transcribed verbatim, and analyzed using the qualitative analysis software NVivo (QSR International, Burlington, MA).^28^ Two primary coders (N.T. and M.C.) performed coding. Rigor in qualitative methods was addressed through the use of Braun & Clarke’s 15-point Checklist of Criteria for Good Thematic Analysis^29^ and Yardley’s Quality Principles.^29^

## Results

Between January 1, 2025, and August 1, 2025, 61 patients were screened for study inclusion, and 48 patients were enrolled. Four patients died before the 4-week assessments and thus were excluded from the analyses. All 44 remaining patients successfully completed the intervention up to the 12-week timepoint of the 6-session intervention delivery and follow up interviews.

The median age was 70 (IQR 63-77), 24 (50%) were male, and 52% (25) had metastatic PDAC, 14 (29.1%) had borderline resectable PDAC, and 9 (18.8%) had resectable PDAC at diagnosis, while 73% had a tumor in the head/neck, 8% were in the body, and 19% in the tail of the pancreas. More than half of the patients (N=32; 67%) were Hispanic, and 65% were English-speaking, while 35% were primarily Spanish-speaking. (see Table 4)

**Table 4.**
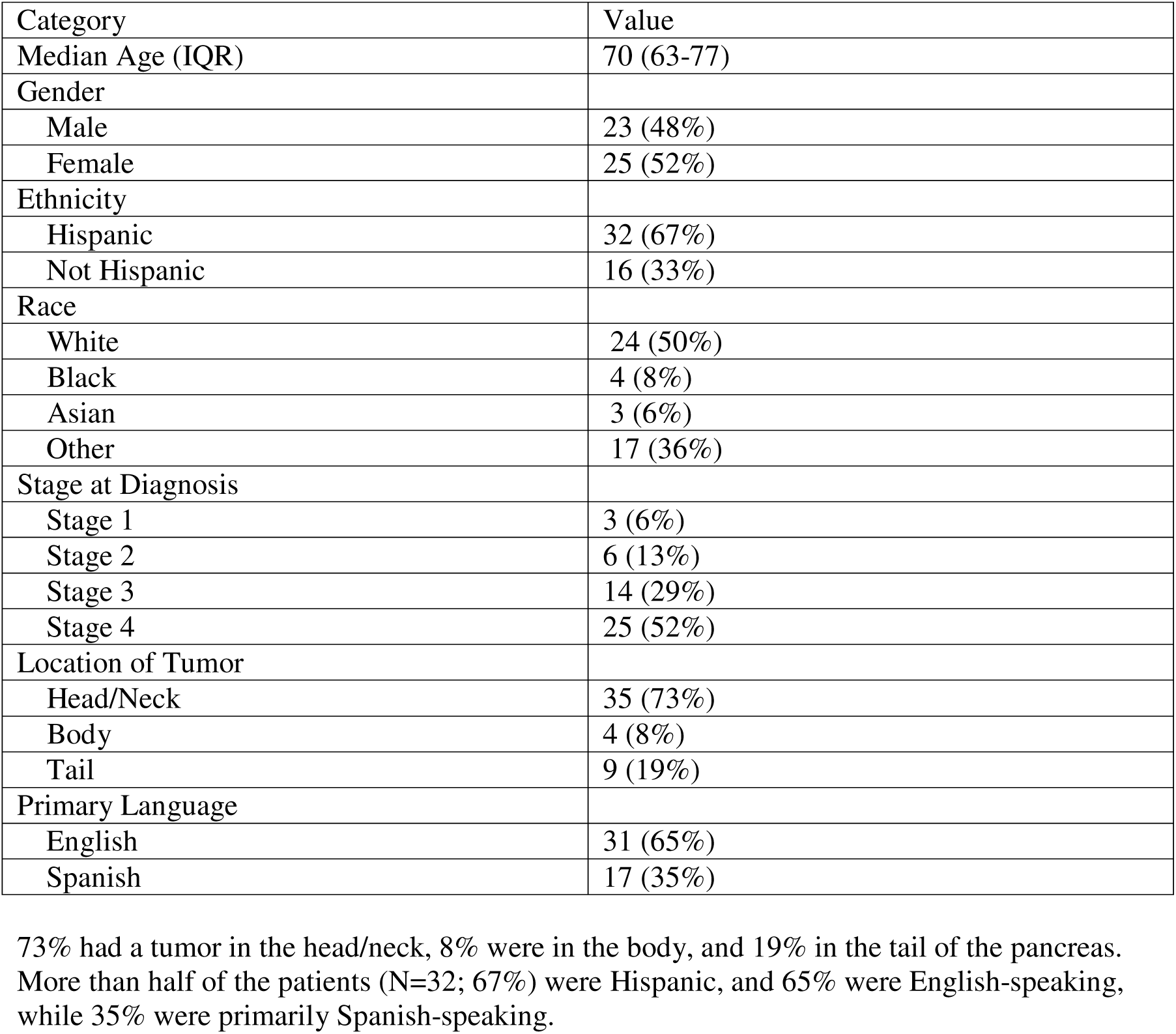
Baseline Characteristics of the Patients in the Study.

### Acceptability of the intervention

Regarding reach, 79% of eligible patients participated in the study, with all participants completing the intervention and completing an exit interview. Regarding effectiveness, the only patients who dropped out were those who had died from PDAC (8% of pts), and the median AIM score was 18 (IQR 14-20), with 94% of patients completely agreeing that the intervention was approved. Regarding adoption, common themes highlighted by patients included the CHW being incredibly helpful to both patients and providers, reducing stress about scheduling appointments, and serving as an essential asset when their symptoms worsened. Regarding implementation, all patients answered at least Twilio SMS ESAS questions, with 84% completing all SMS surveys. Furthermore, participants said the intervention was useful, easy, and consistent with care from their clinical team. (Table 5 for sample quotes from patients and providers) Regarding maintenance, the CHWs reported that the intervention was sustainable and feasible in this population, while physicians did not report additional workload or burden and instead stated that the intervention helped them understand what symptoms to focus on.

**Table 5.**
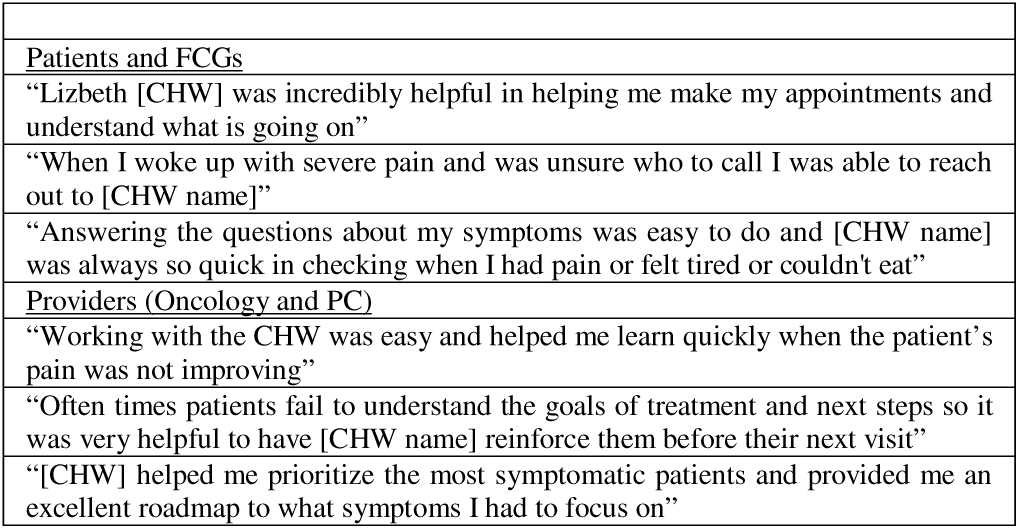
Qualitative Comments from Patients, FCGs and Providers.

### Impact on Quality of Life

The median baseline FACT-Hep total score was 141 (IQR 77 – 182), and it decreased at 4 weeks 127 (69-155) at 12 weeks and to 99 (89 – 121). However, the NFHSI-18 did improve at 4 weeks compared to baseline (55 vs 68, P=0.01)

### Impact on Palliative Care Utilization and ACP

The median PCAS score was 25 (IQR 19 – 29) at baseline (which correlates to Opposed to Palliative Care) but improved to 46 (IQR 37 – 58, p <0.001) at 4 weeks, which correlates to Optimistic. Similarly, the ACP Engagement Survey scores improved from a baseline median of 7 (IQR 4 – 12) to 21 (IQR 14 – 24, p <0.001) at 4 weeks. 40/48 (83.3%) of patients completed ACP, including an advanced directive and POLST form this improved form 4/48 (8.3%) of patients at baseline.

### Impact on Healthcare Utilization

Forty-three subjects (89.5%) utilized telehealth for one of their symptom-focused visits. Thirty-one (65%) of patients saw a nutritionist, while 83.3% completed germline genetic testing. Patients had a median of 2 (1-5) ER visits with 2 (1-4) hospitalizations. Fifty-two percent utilized hospice during the study period.

## Discussion

This pilot feasibility study demonstrated that a CHW-led intervention delivering early connection to stepped PC, care navigation, and disease education was feasible and acceptable for patients with PDAC and their FCGs. Prior studies have demonstrated that CHWs delivering a structured telephone intervention supervised by nurses can increase ACP rates and reduce acute care utilization in patients with advanced solid and hematologic malignancies in Veterans^30^, privately insured patients^19^, Medicare Advantage beneficiaries^31, 32^, and, most recently in low-income and racial and ethnic minorities.^33^ However, in these studies, the health insurance benefit was modified so that it covered PC and behavioral health services, with the removal of prior authorization for oncology services and coverage for CHW services. Our study continues to add evidence that CHWs can play a key role in engaging patients with cancer, including tailoring for PDAC. This intervention supported integrating the CHW into clinic visits with the pancreatology, oncology, and PC teams and functioned with multiple payors without changing current reimbursement models, thus demonstrating efficient care delivery in existing healthcare systems.

Despite evidence that EPC in patients with PDAC improves quality of life, shortages of PC providers, particularly in rural and community settings have limited uptake of PC.^7, 8, 10^ Multiple solutions have been proposed to address this gap in PC providers. One potential solution is a stepped model of PC, in which patients receive a minimum required level of contact with a PC specialist, while those with more severe symptoms have their PC treatment intensified (“stepped up”). Solutions utilizing CHWs have also been studied, focusing primarily on symptom screening and advanced care planning for chronic illness and cancer.^34^ However, no studies with CHWs have been conducted in PDAC, a condition with unique challenges given a substantial symptom burden at diagnosis, with rapid progression of morbidity, and poor overall survival. Our study is novel in that we demonstrated that CHWs can effectively serve as an extension of the PC team, provide symptom self-management to reduce symptom burden, and facilitate digital symptom monitoring and PC escalation for patients whose symptoms worsen. This study also included tailored early intervention for PDAC-specific symptoms, such as biliary and gastric outlet obstruction. Furthermore, with CHW support, ACP rates among PDAC patients at our institution increased substantially from. 24% for to 83%.^35^

Our study is also novel in its use of a simple, SMS-based digital application to assess weekly symptom burden. Prior studies have demonstrated that implementing PROs in Oncology can improve overall survival, reduce acute care utilization, and enhance patients’ quality of life.^36,37^ However, implementing PROs using paper forms can be burdensome, so there has been strong interest in transitioning to electronic collection of routine PROs.^38^ However, electronic PRO collection threatens to introduce digital disparities with substantially lower rates of successful collection in racial and ethnic minorities, older patients, and rural patients.^39^ This has primarily been attributed to digital PRO implementation requiring mobile patient portals and smartphone applications, thereby limiting implementation in patients without smartphones or internet access.

In contrast, in this study, we used feedback from qualitative interviews to implement an SMS-based solution (Twilio) that does not require an app or complicated survey questions for digital ESAS collection. CHW helped set this up for the patients. This resulted in all patients completing PRO collection, and patients completing 85% of weekly surveys, demonstrating that this method was feasible. Furthermore, patients reported that the SMS messages were easy to respond to and took minimal time and effort.

While our study has many strengths, it does have several limitations. The first limitation is that, given the observational nature of this study, we were unable to understand how this CHW-led intervention would compare to usual care. Another limitation is that our study was conducted in a tertiary cancer center with a diverse population (60% Hispanic, 10% Black), with the majority of patients having advanced-stage PDAC. This intervention was provided without cost to patients, but several of them were afraid that this was an expense that might impact their medical insurance limits or access to their treatments. Our results may not apply to other practice settings or populations, though we would argue that, especially for cancers with such poor outcomes, this type of intervention could be especially helpful.

In conclusion, we demonstrated that a novel CHW-led intervention combining digital SMS-based symptom monitoring and educational videos was feasible and acceptable among a diverse cohort of PDAC patients. Patients who received the intervention had reduced symptom burden at 4 weeks, along with improved PC utilization and ACP rates compared with other patients. Further comparative studies are needed to determine whether CHW interventions can reduce morbidity and mortality in PDAC.

## Data Availability

Data produced in the study are not available for use to protect participant confidentiality.

## Abbreviations used in this paper

CI: Confidence Interval
CHW: Community Health Worker
PC: Palliative Care
PDAC: Pancreatic Ductal Adenocarcinoma
RCT: randomized control trial
PRO: Patient-Reported Outcome

